# A mixed-methods multi-site case study of a person-centred intervention for constant observation in hospitals with people living with dementia

**DOI:** 10.1101/2025.03.04.25323366

**Authors:** Melanie Handley, Danai Theodosopoulou, Nicky Taylor, Rebecca Hadley, Claire Surr, Claire Goodman, Rosemary Phillips, Rowan H. Harwood

## Abstract

**Introduction:** Constant observation is widely used with people living with dementia admitted to hospital when identified at risk of harm to themselves or others. Staff allocated to closely monitor individual or small groups of patients intervene when there are safety concerns and may engage with patients’ psychosocial needs. However, care is inconsistent and dependent upon individual and organisational factors. This study aimed to understand whether a co-designed intervention could facilitate person-centred approaches through staff allocation to constant observation.

**Methods:** A mixed-methods multi-site case study explored implementation over 12 weeks in three English hospitals with six wards and one hospital-wide team. Interviews, observations and surveys were analysed using Normalisation Process Theory to explain interactions between individual and organisational contexts, the work of implementation and responses of those involved.

**Findings:** We recruited 153 participants - staff (n=88), people living with dementia (n=71), family supporters (n=4). The intervention was well received and considered useful by staff. Incremental changes, such as staff initiating non-task related conversations with patients and using tools to inform actions for reducing distress, were observed. However, establishing the importance of psychosocial, alongside physical and medical, needs was not achieved. Staff found it difficult to challenge the dominance of medical management and organisations’ priorities to minimise risk. Fears that discussions about constant observation with family supporters might upset them or result in accusations of inadequate care inhibited work to collect and share potentially useful information.

**Conclusion:** The intervention endorsed and supported staff to focus on the quality of their care work; this was not usual practice. Routine use was impacted by prior knowledge of dementia, how the intervention aligned with ward practice and competing priorities. Additional work is required to support the shift from work organised as a reaction to urgent, risky situations to work that supports prevention and enhances care.

## Introduction

For people living with dementia, admission to hospital can be distressing and disorientating. Events, such as falls, arising from misjudgement, disorientation and an unfamiliar physical and psychosocial environment can lead to longer lengths of stay, decreased mobility and reduced independence (1). When staff do not understand behaviours such as getting up or resisting care as a communication of unmet needs they may respond in ways that increase a person’s distress (2). One way of mitigating risks is to allocate specific staff to be with people with dementia to respond to signs of distress and pre-empt harm to themselves or others. This practice is often referred to as constant observation (3–5).

During constant observation, one member of staff is allocated to closely monitor patients either through one-to-one supervision, also known as ‘specialling’, or for a small group of patients, commonly referred to as ‘cohorting’ (Handley et al, 2023). While constant observation is supervised by a senior nurse, direct patient care is often the responsibility of non-registered staff, such as clinical support workers and healthcare assistants. These staff may be temporarily assigned to the ward, have little training in dementia care and not have received instructions for how to provide care beyond keeping someone safe. Incentives to work proactively to reduce distress and respond to individual needs are often outweighed by a preoccupation with risk reduction (6, 7). Staff providing constant observation are likely to favour approaches that can be restrictive and have the unintended consequence of frustrating the person with dementia (8).

Person-centred approaches are widely accepted as best practice for the care of people living with dementia and are recommended in National Institute for Health and Care Excellence (NICE) guidelines (9–11). The practice of constant observation theoretically offers the opportunity for staff involved in care to focus on the individual’s psychosocial needs, with personalised interactions, which might provide further opportunities for preventing distress and risk (12). In theory, staff allocated to constant observation could be enabled to learn more about a person and use that knowledge to inform their own practices and those of their colleagues, but in practice this rarely occurs. Consistent, person-centred care can be difficult to achieve and sustain in hospital settings due to competing demands and priorities (13, 14). The benefits of person-centred approaches have been demonstrated in care homes (15) and empirical evidence for its use in hospitals suggests improved patient experience (16–18).

This paper reports on the implementation of a co-designed intervention devised to change the organisation and delivery of constant observation in ward settings. The CONNECT Enhanced Care intervention was based on best-practice findings from a literature review and prior ethnographic study, and was co-designed with hospital staff, including ward managers and senior nurses, people living with dementia, and family supporters (see below). The intervention aimed to identify, record and share information that would enable staff to anticipate a person’s needs and incorporate what they knew in their approach to care. It comprised four paper-based tools – Making a Difference document (care planning, handover and record of successful interventions), Patient Comfort booklet (patient profile, background and biography), Family and Friends information leaflet, Peer Reflection tool (staff development). These tools linked to social processes, such as handovers and bedside support. The intervention aimed to promote:

i) proactive assessment of needs.
ii) discovery of important personal information.
iii) engagement with family supporters.
iv) information sharing with colleagues.
v) opportunities to debrief and learn from each other.

This study aimed to evaluate the implementation of the CONNECT Enhanced care intervention, using Normalisation Process Theory (19) to structure data collection and analysis. Specifically, we evaluated whether hospital staff could use and embed person-centred approaches with people with dementia during constant observation practices.

## Methods

### Study design

A multi-site, mixed methods evaluation was undertaken between January 2023 – May 2023 in three geographically disparate district general hospitals (19), using a case study design, with each site as a case (n=3) and wards and the hospital-wide team as embedded cases (n=7).

### Recruitment

Hospitals were purposively selected to reflect the different ways constant observation was organised (see table 1). Wards at each site and one hospital-wide team were recruited which regularly used constant observation and committed to testing use of the intervention.

**Table 1:** Predominant use of constant observation across sites.

| Site | Organisation of constant observation |
| --- | --- |
| Site A | Team of non-registered support workers work across the hospital to provide constant observation to people living with dementia. Overseen by more senior non-registered team leads and a senior dementia specialist nurse |
| Site B | Constant observation predominantly allocated to permanent ward staff (clinical support workers) |
| Site C | Mix of permanent ward staff (clinical support workers) and temporary non-clinical staff brought in and funded solely to provide constant observation |

### The intervention

The CONNECT Enhanced Care intervention is a complex intervention that relies on engagement from multiple parties working together. Co-design of the intervention occurred across eight regional online and in-person meetings (n=24), and two cross-site meetings. We purposefully recruited co-designers to represent a range of roles, responsibilities and experiences of constant observation (providing direct care, line management and oversight of staff providing direct care, training) and dementia (living with, supporting someone, working with). A total of 33 people from across the three sites participated between April 2022 and November 2022 including ten senior and experienced clinicians (nurses, allied health professionals, doctor) with managerial and strategic responsibilities for constant observation and care of people living with dementia (see table 2).

**Table 2:** CONNECT Enhanced Care co-design participants by site.

| Site | Staff | People with dementia | Family supporters |
| --- | --- | --- | --- |
| Site A | n=10<br>(1 Clinical support workers, 1 Student Nurse, 2 dementia trained volunteers, 2 Senior Nurses<br>1 Dementia Specialist Nurse<br>2 Allied Health Professionals<br>1 Doctor) | 1 | 5 |
| Site B | n=6<br>(2 Clinical support workers<br>1 Registered Nurse, 2 Senior Nurses, 1 Dementia Specialist Nurse) | - | 1 |
| Site C | n=5<br>(2 Clinical support workers<br>2 Senior Nurses, 1 Dementia Specialist Nurse) | 4 | 1 |
| Total | 21 | 5 | 7 |

Co-designers debated what was needed and what might work, informed by evidence from the first phase of the study that included; i) a systematic review of what supports person-centred constant observation and, ii) mapping of constant observation practices and processes at the three sites. This led to the development of four document-based tools (table 3).

**Table 3:** The purpose and intended use of the CONNECT Enhanced Care Intervention tools.

| Tool* | Purpose | Intended use |
| --- | --- | --- |
| Making a Difference document | Provides a framework for care planning and possible actions during each period of constant observation.<br>To encourage sharing psychosocial information and tips for working with the person during handovers and through written communication. | Referred to and completed by staff providing direct care.<br>Handover of information for working with the person. |
| Patient Comfort booklet | To document and share what staff learn about a person when working with them.<br>Guide conversations with a person to finding out their preferences and needs.<br>Completed sections used to inform conversations, activities, or support when distressed. | Completed by staff with family supporters, person living with dementia and other staff. |
| Family and Friends leaflet | To inform family supporters about constant observation and encourage them to share useful information about the person.<br>To encourage communication between staff and family supporters. | Handed out to family supporters by staff. |
| Peer Reflection tool | To improve practice and learn through structured conversations that share helpful strategies, positive experiences and challenging situations during constant observation. | Brief, in shift reflection sessions between small groups of staff. |

\*Tools are available to download [https://arc-eoe.nihr.ac.uk/news-blogs/news-latest/new-resources-](https://arc-eoe.nihr.ac.uk/news-blogs/news-latest/new-resources-launched-person-centred-approaches-hospitals-people-living) [launched-person-centred-approaches-hospitals-people-living](https://arc-eoe.nihr.ac.uk/news-blogs/news-latest/new-resources-launched-person-centred-approaches-hospitals-people-living) .

The four tools were designed to help staff gather and share information relating to a person’s psychosocial needs and to identify actions to minimise risks in ways that were acceptable and beneficial to the person. Central to the design was that the tools could be incorporated into existing ward-based systems and processes, thereby limiting additional work to staff.

Each ward manager identified two members of staff to act as implementation champions who were supported by the researchers in introducing the tools and the rationale for their use to the rest of the ward team. They received step-by-step guidance on how staff might use the intervention as part of routine work.

Researchers also joined a series of handovers and staff meetings, both online and in-person, to introduce themselves and the study. Information sheets were made available to all ward staff and the hospital-wide team and brief videos explaining the intervention were distributed to staff via WhatsApp and email by managers. Posters about the study were displayed in staff rooms and communal areas. All implementation champions were offered weekly contact with researchers at a mutually agreeable time and place, in-person or online, to support their role and capture feedback on use of the intervention.

At the time of implementation, COVID-19 continued to disrupt hospitals with two study wards (A2 and C2) reporting changes to admissions criteria and ward closures several times during the study period. Additionally, all staff were experiencing the ongoing challenges of having worked through the pandemic, with pressure on hospitals to reduce waiting lists and strikes by various occupational groups. As a result, some planned research activities were restricted, rearranged or delayed.

### Participant Eligibility

All hospital staff working on the study wards or those who worked across the hospital with people living with dementia were eligible to participate. People living with dementia were recruited to the study if they were in receipt of constant observation. Family supporters were eligible if the person living with dementia whom they supported received constant observation during their hospital stay.

### Data Collection

The intervention was used by staff and with people living with dementia allocated to constant observation during a 12-week period.

Data collection about implementation occurred at four time points prior to and during implementation (baseline, week 1/2, week 6/7, week 11/12) (see supplementary file 1).

The evaluation was based on an implementation framework, Normalisation Process Theory (19). Four NPT constructs assess the extent to which: staff understood the purpose and use of the intervention (coherence); the actions different staff undertook to use the intervention (cognitive participation); how staff worked together to realise the benefits of the intervention (collective action); and staff reflections on the extent to which it was possible to embed person-centred approaches into their work (reflective monitoring). Qualitative data were collected in the form of semi-structured interviews based on the NPT constructs and observations of the delivery of constant observation and the intervention in ward areas. Interviews with staff, patients and family carers explored perceptions and use of the intervention. General observations were undertaken to understand the extent to which the intervention was adopted and integrated into daily practice.

Observations took place three times (morning, afternoon and evening) during each period of data collection on each ward. During observation periods, Cohen-Mansfield Agitation Inventory – Observation (CMAI-O)(20) and Quality of Interactions Schedule (QUIS)(21) observational tools were also completed to understand the context of intervention use, these findings will be reported elsewhere.

An adapted Normalisation Measure Development (NoMAD) instrument (22) was self-completed by staff at weeks 6/7 and 11/12 to quantify perceptions of the feasibility and acceptability of the intervention following the initial weeks of implementation. NoMAD was not completed in weeks ½ to reduce data collection burden on staff. The NoMAD instrument comprises 23 questions based on the four NPT constructs and were scored using a 5 point scale of agreement (strongly agree; agree; neither agree nor disagree; disagree; strongly disagree) and summed.

Implementation champions recorded intervention use each week for 12 weeks to understand how the intervention was used and by whom in each of the participating wards and hospital-wide team.

### Ethical approval

Consent to participate by all participants (people living with dementia, family supports and staff) was informed, voluntary and ongoing. If the person living with dementia lacked mental capacity to consent to participate, consultee agreement was obtained from a family carer or friend in accordance with Section 32 of the Mental Capacity Act 2005. Verbal consent was obtained for observations (general and using validated tools) and written consent for all other data collection. Ethics and governance approvals were gained (Yorkshire & The Humber - Bradford Leeds Research Ethics Committee, ref 21/YH/0045).

### Analysis

Throughout data collection, insights and examples were debated in weekly meetings with the core research team (MH, RH, NT, DT) and in monthly full team meetings (MH, CG, RHH, RP, CS, RH, NT, DT). Normalisation Process Theory provided a conceptual framework for analysing the work of implementing interventions (23). The four NPT constructs and their components provided the structure for explaining the work of implementation, investigating how the intervention linked with existing practices, processes and factors within the ward environments, and understanding the effects of using the intervention for staff, patients and family supporters (table 4). Qualitative data transcripts from interviews and observations were imported into NVivo 12 and coded by two researchers (MH, DT). NoMAD data were entered into excel with ratings converted into numbers (‘strongly agree’ = 5, ‘agree’ = 4, ‘neither agree nor disagree’ = 3, ‘disagree’ = 2, ‘strongly disagree’ = 1). Data were imported from excel into SPSS and summarised using descriptive statistics. Differences in responses at week 6/7 and week 11/12 were assessed where participants completed NoMAD at both time points using the Wilcoxon matched-pair signed-rank with 95% confidence intervals.

**Table 4:** Application of the Normalisation Process Theory Constructs.

| NPT construct | Components | Application |
| --- | --- | --- |
| Coherence | Differentiation<br>Communal specification<br>Individual specification<br>Internalisation | Sense-making work for the individual and communal value and benefits implementing the CONNECT Enhanced Care intervention. |
| Cognitive participation | Initiation<br>Enrolment<br>Legitimation<br>Activation | The work staff engaged with to implement the intervention within existing work practices. |
| Collective action | Interactional workability<br>Relational integration<br>Skill set workability<br>Contextual integration | How the work of using the intervention was distributed across teams and the skills required to use the intervention in this way. |
| Reflective monitoring | Systematisation<br>*Communal appraisal<br>Individual appraisal<br>*Reconfiguration | Appraisal of the intervention's usefulness for staff, patients and family supporters and the impact to existing practices. |

\*Evidence for these components was not identified

### Findings

The CONNECT Enhanced Care intervention was tested on two wards at each of three hospitals (n=6 wards) and with one team working across one hospital site providing constant observation for patients with dementia (n=1, total embedded cases n=7) between January and May 2023. A total of 153 participants was recruited (table 5).

**Table 5:** Participant recruitment by site.

|  | Participants |  |  |
| --- | --- | --- | --- |
| Site | Staff | People with dementia* | Family supporters |
| A | 31 | 17 | 2 |
| B | 32 | 21 | 0 |
| C | 25 | 33 | 2 |
| Total | 88 | 71 | 4 |

\*Number includes verbal consent for observations

Four wards were acute medical or geriatric, one was orthopaedic, and one was an older people’s admissions ward. Constant observation for people with dementia was a reported as a model of care regularly used on all wards with estimates that between four and seven patients were being observed on each ward each week. Characteristics of the participating wards are included as a supplementary file (supplementary file 2).

### NoMAD findings

A total of 39 staff across the sites completed a NoMAD instrument. Twenty-nine completed week 6/7 surveys, 30 completed week 11/12 surveys with 20 staff completing both and 19 staff completing only one time point (see supplementary file 3). Responses across the domains demonstrated agreement or strong agreement that the CONNECT Enhanced Care intervention was acceptable and feasible to use, with a median score of 8 out of 10 that staff felt it could become part of their everyday work. However, at week 11/12, there was no change in score for staff feeling the intervention was part of their normal work (median score 6/10).

Normalisation Process Theory constructs were rated positively, with the majority of responses for all components rated ‘agree’ at both time points. The exception was to the statement “The CONNECT Enhanced Care intervention disrupts working relationships” within the collective action construct, the majority disagreed at both time points. There was a trend towards improved mean scores for coherence, cognitive participation and collective action between week 6/7 and week 11/12 although no statistical significance was found with participants who completed surveys at both time points (table 6 and 7).

**Table 6:** NoMAD construct results for weeks 6/7 and 11/12.

|  | Week 6/7 |  |  |  |  |  | Week 11/12 |  |  |  |
| --- | --- | --- | --- | --- | --- | --- | --- | --- | --- | --- |
| Construct | n | Mean<br>(score 1 to 5) | SD | Median | Min/<br>Max | n | Mean<br>(score 1 to 5) | SD | Median | Min/<br>Max |
| Coherence | 26 | 3.9 | 0.4 | 4 | 3/5 | 29 | 4.2 | 0.5 | 4 | 3.3/5 |
| Cognitive participation | 27 | 4.1 | 0.4 | 4 | 3.3/5 | 28 | 4.3 | 0.5 | 4.3 | 3.5/5 |
| Collective action | 25 | 3.6 | 0.3 | 3.8 | 2.9/4.1 | 29 | 3.6 | 0.5 | 3.6 | 2.9/5 |
| Reflexive monitoring | 27 | 4.1 | 0.4 | 4 | 3.5/5 | 29 | 4.2 | 0.5 | 4 | 3.3/5 |

SD: Standard deviation, min/max: minimum / maximum

**Table 7:** Results of week 6/7 and week 11/12 analyses using Wilcoxon matched pair signed rank test.

| Construct | n | Week 6/7 |  |  |  | Week 11/12 |  |  |  | sig |
| --- | --- | --- | --- | --- | --- | --- | --- | --- | --- | --- |
|  |  | Mean | SD | Median | Min /<br>Max | Mean | SD | Median | Min/Max |  |
| Coherence | 17 | 4 | 0.3 | 4 | 3.5 / 5 | 4.2 | 0.5 | 4 | 3.5/5 | p=0.120 |
| Cognitive participation | 18 | 4.1 | 0.4 | 4 | 3.5 / 5 | 4.3 | 0.5 | 4 | 3.5 / 5 | p=0.063 |
| Collective action | 17 | 3.6 | 0.2 | 3.5 | 3.3/4.1 | 3.7 | 0.5 | 3.75 | 2.9/5 | p=0.332 |
| Reflexive monitoring | 18 | 4.1 | 0.4 | 4 | 3.5/5 | 4.2 | 0.5 | 4 | 3.8/5 | p=0.259 |

Sig: significance

### Qualitative findings

The following account is based on interview and observation data.

### Coherence

When introduced to the CONNECT Enhanced Care intervention, staff understood its purpose, were able to articulate how it could be used and how it could be beneficial in their work and the people they were working with. There was an underlying and shared understanding that existing practice was not oriented to person-centred care. The dissonance between current practice and what ought to be done was recognised.

#### Differentiation

During baseline data collection, verbal handovers and informal exchanges between staff regarding patients with dementia allocated to constant observation focused on summaries of medical needs, risks and behaviours likely to lead to harm to the patient or others. There were few examples where staff shared a patient’s emotional and social needs and little evidence that these needs were documented in care records. Staff supporting implementation (ward managers and implementation champions) discussed how the intervention could provide a framework for incorporating psychosocial information as part of their everyday work. Several staff commented that the inclusion of the suggested strategies for working with patients’ different needs and risk-related circumstances provided a framework for extending the range of information considered to systematically improve awareness of different needs and attempting alternative approaches.

> “before this [using the intervention], we were all just doing what’s best for the patient, but this has actually given us a guide to actually do it in the right way, and it’s actually helping to make sure that every need the patient needs has been met. (Nurse, C2_10)”

The components of the intervention that legitimised staff decisions to apply what they knew about a person to their care increased their confidence and valued the skills of the team. It introduced a method that endorsed and supported staff when they focused on the quality of their care work. This was not usual practice.

#### Communal specification

Staff rotas, shift patterns, annual leave, ward closures (due to COVID-19) and sickness meant it was difficult for researchers to inform and train all staff in the use of the intervention, maintain awareness of the study and promote consistent use of the intervention on the wards throughout the 12 weeks. Regular visits from the researchers with the staff responsible for supporting implementation helped to maintain commitment. Meetings with ward managers and implementation champions from wards A1, C2 and team A5 focused on how implementation of the intervention occurred outside of research team visits. However, equivalent meetings with staff in wards A2, B1, B2, C1 were difficult to plan and were often cancelled or interrupted. Researchers perceived that their visits to these wards were often an inconvenience and could lead staff to avoid contact with researchers. During interviews conducted at the end of the 12 weeks, these staff provided explanations for their limited capacity to support use of the intervention, recognising how this impacted use of the intervention by ward staff:

> “Recently we’ve had a lot of difficult things going on in the ward, like our patient group and infections and then sickness, and there’s lots of things that probably prohibited it being more effective.” (Ward Manager, A2_19)

With the exception of wards A1 and B2, who volunteered to be involved following conversations with the research team, all wards were nominated to participate by Senior Management. For at least one ward (C1) recent changes to the ward leadership combined with the ward characteristics meant their capacity and readiness to support implementation was not fully appreciated by Senior Management and only became apparent to researchers once implementation had commenced.

#### Individual specification

Staff allocated to constant observation discussed how the intervention facilitated approaches that could be incorporated into their work monitoring risk and enhance a person’s care. Using the resources meant they were more likely to understand the person’s needs, share that information with colleagues and use what they had learned to adapt care practices to suit the person:

> “I think it is useful because it can make you think a little bit more about the patients and what can be done and what can’t be done to help them. So, you can say ‘we tried something like putting on classical music and that didn’t work, so we tried this one instead.’” (Clinical support worker, A2_03)

These staff also recognised how application of this knowledge could help to form connections with the person, reduce distress and be an opportunity to invite the person’s supporters to have more understanding of the care they were receiving. For other ward staff who were involved with constant observation on the periphery there was less clarity about the value of the intervention for patient care, with the intervention was seen as additional work that was not their responsibility to take on.

#### Internalisation

Contrary to assumptions of the intervention that it could provide continuity and support for staff who were only occasionally involved in constant observation, it had most uptake by those who had experience of doing this work and could see that risk reduction alone was insufficient to support patients well. A prerequisite was an awareness that practice needed to be improved. These staff reported a prior interest in dementia care or having received additional training. They understood the potential for constant observation to be used in a therapeutic way validating their commitment to a person-centred approach. In contrast, staff who reported ad hoc involvement with constant observation were less able to articulate how the intervention benefitted their daily work. Their irregular allocation to constant observation limited their exposure to the intervention, making it difficult to incorporate consistently in their work.

> “I was doing a one-to-one, a couple or three weeks, two weeks ago so then I saw that poster and I was reading it… so I think I applied it straightaway to my patient… But that was weeks ago… it’s not all the time. That’s one of the weaknesses, because it’s not all the time that we can use it [the intervention].” (Clinical support worker, B1_23)

At sites A and C, where constant observation was often provided by staff who were not permanent ward staff, whether temporary staff or staff who provided constant observation to patients across the hospital, this had the unintended consequence of the intervention being seen as someone else’s responsibility. In these circumstances, patient-centred conversations about the people receiving constant observation were less likely to occur.

### Cognitive participation

The relational work of promoting and sustaining use of the intervention was, primarily, the responsibility of the implementation champions. Ward managers decided who took on this role.

#### Initiation

Ward managers and implementation champions responsible for driving use of the intervention with their teams needed to prompt, persuade and enthuse their colleagues to incorporate the intervention as part of their work. How successful they were reflected their status and influence. For example, on two wards (B1, B2) both implementation champions were clinical support workers and appeared to be the only staff who used the intervention. Interviews with them suggested they had talked with colleagues about the intervention and made sure tools were located with other relevant documents. However, they appeared unable to persuade their colleagues to use the intervention. Over half of ward staff interviewed from these wards (n=7 out of 13) reported they either did not know about the intervention or did not realise they could use it. While implementation champions from B1 and B2 reported using the intervention regularly as part of their work and finding the materials helpful, a lack of wider encouragement and support within the wards, particularly from senior staff hindered use of the intervention. In contrast, implementation champions on ward A1 and team A5 reported and were observed to actively engage their colleagues to use the intervention, spending time going through the resources with individuals and in small groups, making suggestions for their use and referring to the resources during their shift. A combination of their motivation, the influence they had with the wider team due to their seniority, and the support they received from senior ward staff helped to maintain the momentum for use of the intervention:

> Discussing experience of acting as CONNECT implementation champion: “[Ward Manager] has been great with it, and so have the Band 6s [senior nurses]. I think, depending on who you’re working with, they will help and support you along each side of it.” (Clinical support worker, implementation champion, A1_01)

#### Enrolment

Along with the hospital-wide team, at least one ward at each site had been involved in co-design of the intervention (n=5). Despite a strong sense of commitment to the co-design process and assertions that the intervention would be useful to staff, this did not guarantee ongoing commitment to the work of implementing the intervention.

> “I say, ‘if you are happy to be an implementation champion, this folder is for you’. I detect a slight intake of breath and see that she feels obligated (from her ward manager and also probably as she’s engaged in the co-design process under her previous ward manager).” (Field notes, C1)

The intervention had been designed to be used across the care team, including with family supporters, but the work of initiating its use was seen as the responsibility of those providing and supervising constant observation. Other staff reported referring to information captured in the Patient Comfort booklet to support their interactions with the person. However, there was limited evidence of the intervention being used or referred to in care planning beyond discussions with staff from team A5.

#### Legitimation

Evidence of use of the intervention beyond written completion of sections of tools was limited. Aspects of the intervention that required staff to interact with other staff or family supporters to share information were reported as being difficult to instigate or were avoided. Routine handovers continued to be dominated with discussions of medical and physical needs. Staff explained that they found it hard to initiate handovers to busy senior staff that focused on information of personal relevance to the patient they were working with. Some staff also found it difficult to engage in conversations about their work with family supporters during their visits. These staff worried that discussing why they had been allocated to constant observation with family members was not appropriate for their paygrade or might provoke a negative response:

> “[Clinical support worker] feels that speaking to relatives is essential but that some staff don’t have confidence to do this. I asked if they felt confident in contacting family, they said maybe if they had the full picture but there is a worry that relatives could misinterpret the reason for contact or might complain. The staff member references she’s ‘only a clinical support worker’, and explains she feels it’s not her place to talk to family, particularly if medical.” (Fieldnotes, Ward A1)

Their concerns included raising expectations for the level of support a person would have throughout their stay on the ward, upsetting family members by discussing distressed behaviours and being blamed for a person reacting in that way. The resource legitimated what staff knew to be important and how they wanted to respond, but this did not legitimate what they did with that information as part of their work in the ward where other priorities took precedence. As a result, visiting family supporters did not know the person was receiving additional support in hospital and were unable to share information about the person that could improve how staff worked with them because they were not talked to.

#### Activation

Implementation champions and senior ward staff described how they made the intervention easily accessible, visible and linked to current practices and systems through assembling and locating the intervention in logical places.

> “My ward manager made up a pack. In the pack they came with the one to fill out, the leaflet and then the one that we fill out. I put the plastic wallet into the nursing notes so that everyone can see.” (Clinical support worker, implementation champion, B1_13)

This activity alone was insufficient to encourage use of the intervention. However, when these strategies were combined with encouragement, such as reminders to use the intervention or directly handing out the intervention to staff, use of the intervention was more likely. Two senior staff spoke about using the intervention to guide and supervise staff providing constant observation by checking what staff knew about the person they were working with and helping them to think through how they would use this information to enhance the person’s care.

> [discussing use of the intervention] “When I’m doing my rounds, I will always ask ‘Where is this assessment and care plan? What are you doing about this? What is your involvement into this? Have you engaged the family?’.” (Senior nurse, Site A)

These senior staff discussed how the intervention complemented their work and were able to easily incorporate it as part of their current practices.

### Collective action

Underlying collective action was the value that staff placed on constant observation, whether they recognised the potential for it to be a therapeutic activity, and if psychosocial information was viewed as important.

#### Interactional workability

Clinical support workers and staff temporarily assigned to the ward to provide constant observation found it difficult to use the intervention with their colleagues unless senior staff facilitated opportunities.

> “…that [Making a Difference document] was very useful, because sometimes I don’t get a single bit of information about my patient… I’ve got to approach a nurse. They just are more busy about getting into their handover and they go, oh, yeah, you’re looking after bed so- and-so, and they just give me a number.” (Temporary Constant Observation Staff, C1_15)

Those providing constant observation were reluctant to share information about a person unless it was related to immediate risk or medical concerns. This prioritisation of medical and behavioural information was reinforced through routines on the ward, such as the structure of morning and evening handovers. Personal information and strategies used to reassure and comfort distressed patients, if discussed, were part of separate handovers with senior staff who had specific responsibilities in overseeing constant observation and allocation. For staff, this underlined how clinical care and person-centred care were separate activities.

#### Relational integration

Staffs’ sense of confidence in working with people living with dementia identified as at risk of harm to themselves or others was highlighted through how they used the intervention. Staff discussed their reluctance to use the intervention in a way that identified them as accountable or exposed their work, for example by writing down their observations of how a person responded in different situations. This meant that strategies that had been attempted, regardless of whether or not they were beneficial to the person, were not documented:

> “We discussed the reason for constant observation. [Patient] likes to walk around ward but is not waiting for support from staff. The team lead explained that reassuring the patient that staff are there to provide support with walking did not always work. The team lead noted Patient Comfort booklet had not been used to communicate this as they not found something that has worked.” (Field note, Team A5)

Staff did not document potentially informative patterns of behaviour when constant observation was first initiated, instead stating that patients needed time to settle into the routine of the ward before committing to writing down observations. This observation of the typical patient trajectory for those receiving constant observation impacted the learning and care planning potential of the intervention at a time when it was critical that a person was supported in ways that accommodated and understood their needs. Staff explained not recording this information as they considered it would quickly become obsolete. Instead, staff used the tools during quieter times and when they felt confident that the information recorded aligned with other care partners’ understanding and observations.

> “The nurse explained patient was in a good frame of mind to talk so she sat down and had a chat with him and was able to initiate conversation and add to the booklet.” (Field note, Ward A1)

Of itself, use of the intervention to structure conversations with a patient with dementia was considered helpful to staff and beneficial to patients.

> “It started conversation starters, which then built a relationship with my patients.” (Clinical support worker, implementation champion, B1_13).

#### Skill set workability

The intention of the intervention was to build a picture of a person across the duration of them needing constant observation to support care and care planning. However, there was no evidence the intervention was used in this way; tools were completed at one time point and were not added to. In part, this was linked to how the credibility of what was documented would be established:

> Participant 1: “The problem is, if I fill in a little bit of it, and then you fill in a little and then [colleague] puts in a little bit, then the ward staff put in a bit and then the family…”
>
> Participant 2: “… and put your name.”
>
> Participant 1: “But what’s the point of putting our [name down]…”
>
> Participant 2: “it’s just like a collective… so the reliability of who puts it in.” (Joint interview with clinical support workers, Site A)

Similar discussions related to whether information about a person was representative of their typical way of being. This demonstrated a tension for staff between recording what they observed or learnt about the person through conversations and interactions with them and what those who knew the person well reported. The intervention was creating a different way of recording behaviours that did not align with established practice for documenting patient experiences.

#### Contextual integration

Often the staff providing constant observation did not have access or had restricted access to the electronic notes systems. This limited what they were able to view and update meaning their knowledge of the person relied on what they were told by other staff, what they learnt during their work and impacted continuity of information available to others providing constant observation. Staff temporarily assigned to the ward felt the intervention was beneficial to themselves and the ward team, providing continuity across those involved with the person’s care. However, some ward staff considered introducing the intervention to temporary staff was time consuming and of limited value because of their transient involvement.

> “I think ward staff aren’t thinking about them [temporary staff] using it [intervention]. But it’s really important that they are, because they’re the ones that are providing that enhanced care [constant observation].” (Clinical support worker, implementation champion, C2_01)

This was despite some observed and reported consistency in the temporary staff who worked on the wards and provided constant observation.

### Reflexive monitoring

Apart from the earlier example of a senior staff member using the intervention to guide patient care with their staff, there was limited evidence related to reflexive monitoring of the intervention. Interviews with ward managers, implementation champions and staff on the wards who had used the intervention focused on the need for more training to encourage use of the intervention.

#### Systemisation

A number of staff reflected on their experiences of using the intervention, commenting on the benefits of the tools to themselves and for the patients they worked with.

> [discussing patient with limited verbal communication abilities and who is in isolation for COVID-19] “for this patient, having the Comfort booklet has been very useful for the staff to get to know her and how to reassure her… staff have been able to include the name of the son when they talk to her… she likes to sit quietly and listen to the radio, so the staff put the radio next to her and that’s keeping her happy and calm.” (Clinical support worker, implementation champion, B2_12)

They reported that the Patient Comfort booklet was the most used. The similarity of this tool in terms of gathering personally important information to existing patient profile tools meant staff were able to transfer current practices to this component of the intervention. However, this meant that the ambition for the document to be a dynamic record of staff observations that gleaned useful information about what did and did not work for the person was not achieved.

#### Individual appraisal

Interviews suggested most staff narrowly interpreted ‘use’ as completing the tools rather than also including times when resources were referred to for informing their own or colleagues’ work. The potential of the intervention to encourage a shared sense of responsibility for constant observation was not recognised. However, staff who had referred to tools’ content but not added to them reported that information had been useful for starting conversations with patients, particularly if they were beginning to become distressed by their surroundings or provided tips to comfort them at times of increasing distress.

> “On a nightshift a couple of weeks ago, there was a patient who had Alzheimer’s, and they kept on getting a little bit agitated. And then that’s when I’d refer to the documents and have a quick look through to see if there’s any information that can benefit me, and in making sure that they’re okay, and making sure that everyone around them isn’t then worried about that patient.” (Clinical support worker, C1_04)

This suggests evidence of incremental changes triggered in situations when other help might not have been available.

## Discussion

This study aimed to understand what is needed to enable and normalise person-centred approaches with people with dementia in hospitals during constant observation practices. The CONNECT Enhanced Care intervention required staff to share responsibility for its use in a hard-pressed environment where constant observation was often delegated to non-registered staff tasked with prioritising the minimisation of risk. The intervention was, for the most part, positively appraised by hospital staff in that they understood its purpose, they believed they would be able to incorporate it into their daily work and they considered, when used, it would be beneficial to patients and family supporters. However, there were tensions between acceptance and welcoming of the intervention at a theoretical level and what appeared to be insurmountable barriers at the practical implementation level. Routine use of the intervention beyond form filling was affected by the seniority of the staff using it, prior experience of dementia care, if staff were part of the ward team, how it aligned with existing methods of recording and responding to patient needs and competing priorities. Many of these factors have been reported as general barriers to patient-centred, relational care across hospital ward settings with people living with dementia and older people (24–26). Specific factors inhibiting use of the intervention in this study were anxieties about how documented evidence might be used and interpreted beyond the purpose of recording events, and the potential of the intervention to reinforce a pre-existing separation of care responsibilities for patients allocated constant observation.

There was some evidence that the intervention could be used as a way of holding the uncertainty and unease of risk management work when used as a tool to agree acceptable actions for care and share responsibility for those actions between a senior team member and staff providing direct constant observation care. Using the intervention in this way helped staff providing constant observation to reconcile deviation from standard practices and provide care that could better meet the person’s needs. It was a conduit for valuing care in an environment where there were few opportunities to recognise and legitimise good practice. This more proactive care was endorsed through the support of the senior colleague who held accountability for this decision. However, most wards did not use the intervention in this way, instead relying solely on staff providing constant observation to complete the tools and individual discretion about whether and how that information was used in their practice. To expect non-registered staff to work creatively and autonomously to promote choice and demonstrate understanding for individual patients who, for the most part, were considered disruptive to getting the ‘real’ and non-negotiable work of the ward done, requires the active support of senior staff and a favourable organisational culture (6, 27–29). For many clinical support workers involved with constant observation in this study, this support, while recognised as important, was not possible to accommodate within the working practices of most wards. As Scales et al. (7) acknowledged, attempts to empower staff must recognise both their relational and positional status and the structural constraints imposed through the norms, processes and cultures of their work.

While all staff recognised the role of family supporters in understanding how to adapt care for the person, fears related to the reactions from relatives and friends related to discussing a person’s safety needs, particularly when related to negative physical and emotional expressions of distress, meant staff often avoided initiating discussions about constant observation with family supporters. Similar to other hospital studies exploring communication between hospital staff and family supporters around care decisions (30, 31), this study found that clinical support workers lacked a framework for these discussions. Specific concerns focused on saying the ‘wrong thing’ and having their practices scrutinised, concerns which have been identified in other studies (32, 33). For some staff, this anxiety prevented them using the intervention with family supporters. While senior nurses stated they encouraged staff, including clinical support workers, to speak with family supporters to find out more about a person, we found no evidence for how staff were prepared or assisted to undertake these potentially sensitive conversations. Further work is necessary to understand reasonable expectations for care related conversations between staff providing constant observation and family supporters and how senior team members are enabled to support this.

The intervention required staff to maintain a dynamic record of psychosocial information and apply this to alleviate distress, reduce risk or support occupation. Evidence showed that staff used the tools differently depending on a person’s level of distress and the length of time since their admission. The intervention was only used when staff felt the person had ‘settled into’ the ward, indicating staff recognised that agitation, distress and potentially unsafe behaviours were common around times of transition, such as admission to hospital and changes of ward location, and that these often spontaneously resolved without specific intervention. A different approach to constant observation, and information needs to support it, may be required specifically at this time. While recognising the distressing nature of being admitted to hospital when unwell, potentially in pain or having delirium and living with dementia, this observation had implications for actions for care. Initial priorities for safety and containment could overlook the exploration of patterns and the use of proactive and potentially therapeutic approaches for people allocated constant observation. Socialised care futility has been used to explain how the collective acceptance of behaviours that do not respond to staff intervention, such as repetitive calling out, justify subsequent staff (non)responses in the context of people living with dementia admitted to hospital (34). Similarly in this study, staff initiation of the intervention with patients with dementia allocated constant observation suggested a shared sense of futility in early assessments and attempts to understand a person’s needs when they were identified at most risk of harm. This belief appeared to be reinforced and rationalised by staff with limited control over their practical capacity to engage with disruption to ward routines yet were accountability for patient safety. To embed care practices that reduce the length of time people are distressed and present as at high risk of harm to themselves or others, these beliefs and constraining factors will need to be challenged.

Building a shared sense of responsibility for risk management activities, such as falls prevention, is recognised as important but challenging when aspects of care are assigned to specific staff groups (35), and can constrain how staff, such as clinical support workers, address the needs of patients. Where the work was allocated to staff who were not part of the ward team this reinforced beliefs that the intervention was not linked to ward or nursing work and was a distraction. This reallocation of care work without active senior oversight raises questions about whether this benefits the service more than the patients (36, 37). In most wards, the intervention reinforced dementia care as separate and disaggregated activity from what other staff do. The nature of constant observation as care provided by non-registered staff, often from outside of the core ward team, exposed the implicit hierarchy of what work matters and how that inadvertently disadvantaged individual patients and their needs. While constant observation has been implemented in hospitals to minimise adverse events arising with patients identified at risk of harm, this has negatively impacted the sense of collective purpose and responsibility for people living with dementia.

Despite considerable co-design with a view to implementation, and theoretical support for the approach and intervention, implementation in practice was, at best, only partly successful. Translating the enthusiasm from the co-design of the intervention to implementation was challenging. Co-design meetings and activities contained staffs’ responsibilities to sharing experiences, ideas and critiquing drafts of the intervention within a specified and restricted time. To successfully implement the intervention, a more intense level of commitment was required often outside of visits from researchers. This was not always possible or prioritised when faced with other demands. The research process was new to some staff and was recognised as more formalised and time consuming than quality improvement efforts they were familiar with. Alternative approaches to implementation, such as Plan Do Study Act cycles, may have been more supportive of implementation efforts. However, it is unlikely an alternative approach would have addressed more complex challenges around ward culture or staff concerns around working with families (38).

### Strengths and limitations

The study was conducted in three regions of England in hospitals that differed in their processes and practices for constant observation with people living with dementia. Feasibility testing using an embedded case study design provided a detailed exploration of enabling and inhibiting factors for implementation within and across settings. Researchers were trained in data collection methods supporting the reliability of findings. However, the NoMAD tool was limited in its contribution to understanding acceptability and feasibility of the intervention and was off putting for staff. This may also have impacted implementation efforts for those staff who found it difficult to separate the purpose of research data collection materials with the intervention tools.

## Conclusion

Hospital staff providing constant observation for people living with dementia are potentially well-placed to improve the quality of care these patients receive, however there are consequences of creating parallel systems of care in a ward setting. With limited authority, staff providing constant observation rely on encouragement and validation from senior staff to embed new practices within the team. While the CONNECT Enhanced Care intervention was acceptable to staff and considered feasible to use within the routines of the ward, additional work is required to support the conceptual shift from work organised as a reaction to urgent, risky situations to work that supports prevention of distress and enhances care experiences.

## Data Availability

All relevant data are within the manuscript and its Supporting Information files.

## Acknowledgements

We would like to thank the all the participants for their involvement. We would also like to express our gratitude to the CONNECT advisory group who provided oversight and guidance for the conduct of the study.

## Supplementary Files

Supplementary File 1: Table 1: Data collection schedule

Supplementary File 2: Table 1: Characteristics of participating wards

Supplementary File 3: Table 1: NoMAD Survey findings at timepoints 2 and 3

